# Behavioural and Pharmacological Interventions for the Management of Pain Perceptions in Rheumatoid Arthritis: A systematic-review and meta-analysis

**DOI:** 10.1101/2025.08.14.25333667

**Authors:** Charlie Joseph Neale, Andrew Soundy

## Abstract

**Objectives:** To evaluate and compare the individual therapeutic efficacy of NICE-recommended physical activity (PA) and pharmacological interventions on pain amongst adults with rheumatoid arthritis (RA).

**Methods:** A systematic-review and meta-analysis of studies published between March 1988 and April 2025 was conducted across seven databases; AMED, MEDLINE, CINAHL Plus, SPORTDiscus, EMBASE, Google Scholar, Web of Science, and reference lists. Included were monotherapeutic randomised controlled trials (RCTs) of DMARDs, NSAIDS, analgesics, aerobic and/or resistance training for managing pain perceptions; measured as change in pre-and-post-intervention scores using the visual analogue scale (VAS). Participants were aged ≥18 years whose condition met American College of Rheumatology (ACR; 1987/2010) RA-criteria. Pooled meta-analyses results were presented as mean differences (MDs) and 95% confidence-intervals (95% CIs). Risk of bias (ROB) and certainty of evidence were assessed with the ROB 2 tool and Grading of Recommendations Assessment, Development and Evaluation (GRADE) approach.

**Results:** Searches identified 3286 articles. 25 trials were selected for inclusion (6468 participants); 14 RCTs of 11 aerobic-and/or-resistance-training programs (n=916), three yoga regimes, an individual joint-protection programme, a trial of Rocabado exercises, and 11 RCTs of 21 DMARD/NSAID monotherapies (n=5552); baricitinib, celecoxib, filgotinib, hydroxychloroquine, ketoprofen, leflunomide(n=2), methotrexate, naproxen (n=2), sarilumab, sulphasalazine, tofacitinib, and upadacitinib. Weighted mean differences in pain perceptions for behavioural and pharmacological interventions were −2.47mm (95% CI: −3.14 – −1.81, p<0.00001) and −11.20mm (95% CI: −11.35 – −11.05, p<0.00001) respectively.

**Conclusion:** Despite inconsistent control of medication histories and PA-prescription, adherence to behavioural and pharmacological interventions can successfully alleviate pain. First-line management using DMARDs or NSAIDs appears to be more effective than yoga, Rocabado exercises, or aerobic and/or resistance training alone.

**Systematic review registration number:** CRD420251069339

**Key Messages:**

- Both pharmacological and physical activity interventions can successfully reduce pain perceptions amongst patients with RA.
- Independent use of DMARDs or NSAIDs appears to alleviate pain more than aerobic and/or resistance-training.

## Introduction

Rheumatoid arthritis (RA) presents a significant global health challenge, particularly within the United Kingdom (UK) where it currently affects up to 1% of the population daily at an enormous annual cost (£4.8 billion) [1, 2]. As we head towards 2050, worldwide prevalence is expected to continue to increase and it is therefore critical for us to accurately support those currently living with condition and prepare ourselves in advance of this anticipated mass rise in incidence [3].

RA is an autoimmune, polyarticular, inflammatory condition of the synovial joints believed to be the result of maladaptive, innate immune system responses [4–6]. This develops at any age, typically peaking amongst 30–50-year-olds, with a greater prevalence amongst females due to an epigenetic-interaction between sex hormones and production of pro-inflammatory cells [1, 7–9]. Given where it manifests itself within the body, RA has a profound effect on the musculoskeletal system that can result in articular and extra-articular symptoms which can fluctuate in intensity due to known heterogeneity in RA-pathogenesis and presentation [10–12]. Widespread joint pain is one of the most commonly reported symptoms of RA [13], thought to be a multidimensional experience as a consequence of reduced neurone sensitivity due to sustained activation of the central and peripheral nervous systems [14, 15], peripheral hyperactivity driven by constant nociception [14], and/or skeletal and soft-tissue damage [16]. Unfortunately, like the majority of symptoms, this pain infamously makes movement difficult and impairs physical function, ultimately leading to a rapidly progressive level of disability and notorious decline in quality-of-life [17]. Finding ways to effectively alleviate this is said to be a top-priority for healthcare professionals and patients alike.

Continuous evolution in our pathophysiological knowledge of RA have enabled the creation of multiple treatment options to improve prognosis. Most-frequently a ‘treat-to-target’ approach is taken to supporting patients involving regular assessment and proactive responses through the type of treatment provided [18]. Current UK-based best-practice guidance for the management of RA amongst adults recommends the use of pharmacological interventions such as conventional disease-modifying anti-rheumatic drugs (cDMARDs) and glucocorticoids, followed if unsuccessful by biological (bDMARDs), targeted-synthetic (tsDMARDs), non-steroidal anti-inflammatory drugs (NSAIDs), and/or analgesics [18]. Each of these classes can play a key role in modifying the disease-process (summarized in Supplementary Table S1) and has been shown to possess strong clinical efficacy [12, 19]. Unfortunately, however, regular use of such medications also carries possible adverse side effects by simultaneously weakening the body’s natural lines of defence [20; pp.20-65]. Equally, heterogeneity in RA also means that not every individual will benefit from every drug, where attempted match-making to individual patient needs has yet to sufficiently meet population demand [21].

In spite of historical clinical misconceptions, NICE [18], The World Health Organization [22] and global governing bodies have suggested encouraging regular physical activity (PA) as an alternative behavioural intervention to assist with this disparity in management. Whilst others such as diet and psychological techniques have received attention, these are not formally-recognized due to lacking robust evidence and therefore were not included in this review. PA possesses a wealth of universal physiological and psychological benefits [23–25]. As for RA, aerobic and/or resistance-training can significantly alleviate pain and contribute to the reduction in intensity of other inhibitory symptoms [26–29], considered to be the result of an internal anti-inflammatory effect through suppression of pro-inflammatory cellular sources (e.g. IL-6, IL-10, IL-1)[30, 31]. However, whilst evidence does exist supporting its use, previous systematic reviews into the efficacy of PA have often been of low-to-moderate methodological-quality, have not included meta-analysis, or not directly compared outcomes with those from pharmacological interventions [27, 29].

Unquestionably, RA places a significant burden on the quality-of-life of patient’s physically, psychologically, and socially. An articular manifestation means that many with the condition face lifelong pain which stands amongst the most-prevalent symptoms experienced. To combat this, it is crucial for healthcare professionals and patients to seek the management plan that can most-effectively help to control disease-activity and meet unique clinical needs. Best-practice guidance currently supports the evidence-based use of both pharmacological and PA interventions but fails to include key research details for credibility and recognise which, if either of these interventions, should take priority within the structure of plans, raising the question as towards which efforts should be made. In hoping to address the limitations of previous research and provide clarity on potential treatment options, this systematic-review and meta-analysis therefore aimed to evaluate and compare the individual therapeutic efficacy of latest-recommended PA and pharmacological interventions on joint pain perceptions amongst adults with diagnosed RA. Our research question asked was; Are behavioural interventions (PA) equally or more effective for alleviating joint pain in RA patients than alternatively available pharmacological approaches?

## Methods

This systematic-review and meta-analysis was performed in line with recommendations by the Cochrane Collaboration [32] and the Preferred Reporting Items for Systematic Review and Meta-Analysis (PRISMA) guidelines (Supplementary Table S2 and S3)[33]. The study protocol was defined and subsequently registered in-advance with PROSPERO (CRD420251069339). Ethical approval was not required as all of the data used was already publicly available and not of any confidential nature to participants.

### Eligibility Criteria

Eligibility of studies for inclusion within this review was assessed according to a PICOS strategy (population, intervention, comparator, outcome, and study design). Table 1 provides a full summary of the PICOS criteria used.

**Table 1.** PICOS strategy criteria for inclusion and exclusion of studies. **Alt text:** Table describing the exact methodological inclusion and exclusion criteria for studies to be considered within this review, separated into population details, interventional details, required comparators, essential outcome measures, ideal randomized-controlled trial study design, and other subject-specific features.

|  | Inclusion Criteria |
| --- | --- |
| **Population** | Adults aged 18 years old and over who had previously received a formal diagnosis of RA in line with the ACR-EULAR Revised 1987/2010 Classifications for RA [34, 35].<br>Studies were excluded if participant's co-morbidities were uncontrolled or where these were specifically investigated (risk of confounding effects). |
| **Intervention** | Any of the following pharmacological or behavioural interventions included within the latest set of best-practice recommendations by NICE [18];<br>**Pharmacological;**<br>• cDMARDs: Methotrexate, Leflunomide, Sulfasalazine, or Hydroxychloroquine.<br>• bDMARDs: Adalimumab, Etanercept, Infliximab, Certolizumab Pegol, Golimumab, Sarilumab, Tocilizumab, Abatacept, or Rituximab.<br>• tsDMARDs: Tofacitinib, Baricitinib, upadacitinib, or Filgotinib.<br>• NSAIDs: Ibuprofen, Diclofenac, Ketoprofen, Indomethacin, Naproxen, Etoricoxib, Celecoxib, Meloxicam, Nabumetone, or Piroxicam.<br>• Analgesics: Paracetamol, Co-codamol, or Tramadol.<br>**Behavioural;** |
|  | • Aerobic and/or Resistance Training.<br>• Physiotherapy (to manage functional impairments, and enhance flexibility and muscle strength).<br>• Exercise programmes for stretching and strengthening of the hand or wrist.<br>• Complementary therapies.<br>Due to potential confounding whilst the efficacy of individual interventions are determined, complex and multifactorial interventions were excluded. Studies were also excluded if they included uncontrolled, concurrent use of background pharmacological and/or behavioural interventions. |
| **Comparator** | Studies were required to include a control-group, placebo, standard care intervention, or non-active alternative that were compared with experimental-groups. |
| **Outcome** | Pain needed to be assessed as an outcome measure using the visual analogue scale (VAS); in cm or mm. In the absence of a gold-standard for measuring pain-perceptions, no other tool is used as consistently [36]. The VAS pain scale has also been demonstrated to possess sensitivity to clinically-important differences, strong construct validity (0.62-0.91) and high-reliability for patients amongst with inflammatory arthritis ($r=0.59-0.94$)[37, 38].<br>Studies must have reported results as a mean difference without or without standard deviation or have made available pre-and-post-intervention scores to have allowed differences between assessment to be calculated. |
| **Study Design** | Studies were limited to include only RCTs. Randomisation minimises bias and confounding variables, allowing for reliable estimates of causality and internal validity of results [39]. |
| **Other Limits** | A full-text version in the English-language needed to be available for review. Translators such as Google Scholar may have been able to support with including texts in alternate languages, however accuracy is often variable depending upon the original language that the articles was published [40] and reliability is shown to be mixed for procedural texts like RCTs [41].<br>Studies needed to be published between March 1988 and April 2025 given that participants were required to meet revised ACR criteria not published until 1988. |
| **Abbreviations:** cDMARDs, conventional disease-modifying anti-rheumatic drugs; bDMARDs, biological disease-modifying anti-rheumatic drugs; targeted-synthetic disease-modifying anti-rheumatic drugs; NSAIDs, non-steroidal anti-inflammatory drugs; NRS, numerical rating scale; VAS, visual analogue scale; RCTs, randomised-controlled trials. |  |

### Search Strategy

Initially, a single reviewer (CJN) independently searched electronic databases for studies published between March 1988 and April 2025. A second reviewer (AS) then replicated the search strategy using Covidence software [42] to account for any articles missed/previously overlooked. Based on previous reviews and suggestions, these databases were AMED, MEDLINE, CINAHL Plus, SPORTDiscus, EMBASE, Google Scholar and Web of Science [43–45]. A manual search of reference lists of included studies and NICE evidence reviews for DMARDs and analgesics was also performed to identify potentially-relevant articles [46, 47]. Searches were defined using database specific subject language and all appropriate synonyms for the areas of interest, combined by Boolean operators, and guided by the Peer Review of Electronic Search Strategies (PRESS) statement table [48]. A copy of the initial search worksheet and full search strategy are available within the supplementary materials of this document (Supplementary Data S1 and S2). References for all included studies were entered into Endnote Online reference management software [49].

### Study Selection

Studies were screened via their titles and abstract against inclusion and exclusion criteria. Full-texts of potentially relevant studies were then reviewed independently by two reviewers (CJN and AS). Only studies receiving a consensus of votes from both reviewers were selected for inclusion.

### Data Extraction

All data was extracted and captured in a bespoke Microsoft Word document by a single reviewer (CJN). For studies deemed eligible, where available, data was collected according to the authors, date of publication, study country, sample size, demographical information (age, gender distribution, disease duration, and disease activity), details of the intervention and comparator, and results relating to the assessment of pain. In the scenario where a study had used repeated measures of assessment, only data for the longest timepoint of intervention was captured for analyses.

### Risk of Bias Assessment

To assess the risk of bias, the Risk of Bias Tool 2 (ROB2) by Cochrane was applied to the individual included studies [50]. The ROB2 tool enables an in-depth analysis of study content across five domains; randomisation process, deviation from intended interventions, measurement of outcomes, missing outcome data and selection of reported results, producing an assigned rating for each study.

### Data Synthesis

Once captured, all data required for synthesis was manually inputted to a customized Microsoft Excel table and transferred electronically for meta-analyses. The therapeutic efficacy of each intervention was assessed according to the magnitude of change in mean difference (MD) and standard deviation between pre-and-post-intervention VAS-pain scores, where a minimally clinically important difference (MCID) was defined as ≥10-15mm [51, 52]. Differences in weighted therapeutic effect were presented with 95% confidence intervals (95% CI), used to calculate effect sizes which were pooled and subsequently compared with that of comparator-groups. A p<0.05 was considered statistically significant.

### Statistical Analysis and Certainty of Evidence

All data and statistical analysis was performed using Microsoft Excel [53] and RevMan software (version 5.4.1) [54]. Any necessary missing data such as standard deviation and change scores were calculated using that which were available and formulae outlined within the Cochrane Handbook [55] and/or published by Hozo, Djulbegovic, and Hozo (2005)[56] or Wan et al.(2014)[57]. As all studies possessed similar characteristics, a uni-directional, uni-variate meta-analysis of the included studies was performed using a random-effects model of pooled data to account for heterogeneity. Pooled mean differences and 95% confidence intervals were calculated on RevMan (Version 5.4.1), represented in forest plots. Heterogeneity was assessed using the I-squared (I^2^) statistic; where values of 0%-40%, 30%-60%, 50%-90%, and 75%-100% were used to reflect unimportant, moderate, substantial, and considerable differences [58], and if discovered, robustness and sensitivity assessed according to sub-group comparisons by gender, type of medication, and exercise-modality. Quality of clinical evidence was assessed with the Grading of Recommendations Assessment, Development and Evaluation (GRADE) approach where pooled data was rated high, moderate, low or very low [59].

## Results

### Study Selection

The full search protocol for this review is described below in Figure 1. Initial database searches (n=2901) and manual screening of reference lists (n=385) identified 3286 relevant articles. Of these, the full-texts of 436 articles were selected for review against inclusion criteria, and 2875 were excluded. In total, 26 studies were chosen to be included within this systematic review and meta-analysis [60–84]. Excluded studies were those that did not meet the inclusion criteria and/or were not appropriate for reasons described in Figure 1.

**Figure 1.**
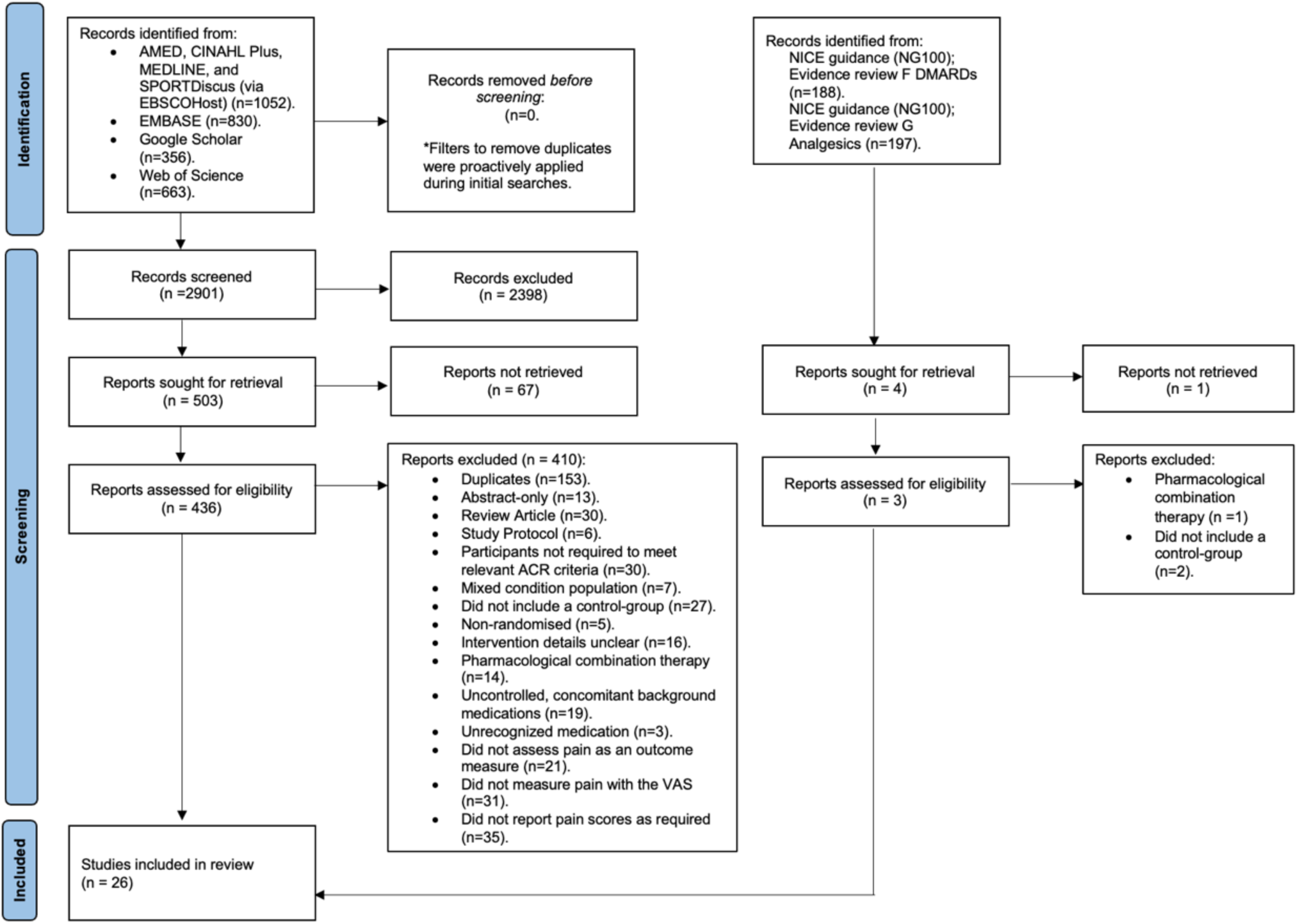
PRISMA flow diagram of search strategy. **Alt text:** Flow chart detailing the web-based databases searched, and number of studies included and excluded at each stage of the search protocol.

### Study Characteristics

#### Behavioural Studies

A total of 14 RCT’s involving 14 interventional exercise programs were included in this review (Table 2); three involving aerobic training [60–62], three including yoga [63–65], two resistance training [66, 67], two combined aerobic and resistance training [68, 69], two bilateral resistance training programs of the hand-and-wrist [70 71], a trial of maxillofacial Rocabado exercises [72] and a community physiotherapy initiative [73]. Studies were conducted across 12 countries; Brazil, Canada, Croatia, Denmark, Finland, India, Ireland, Iran, New Zealand, Spain, Sweden (n=2), Turkey, as well as multi-nationally, with a duration between 6 and 104 weeks. Participants (n=916) included 455 indivduals in intervention groups and 461 in control groups. Age ranged from 47 to 65 years (mean: IG = 53.4, CG = 53.6) where sex distribution and diease duration ranged between 53%-100% female and 3-16 years respectively (mean: IG = 9.2, CG = 9.0).

**Table 2.**
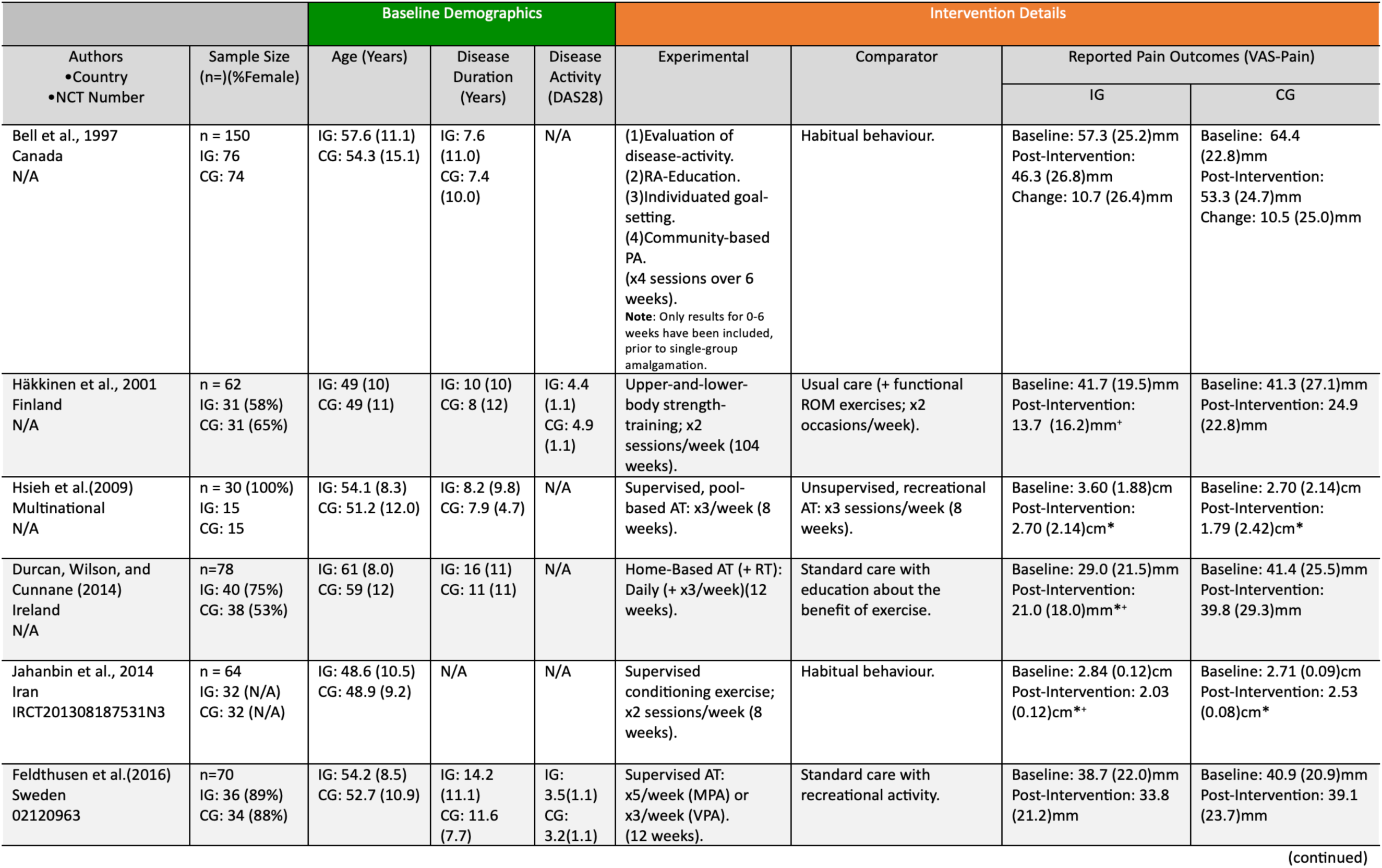
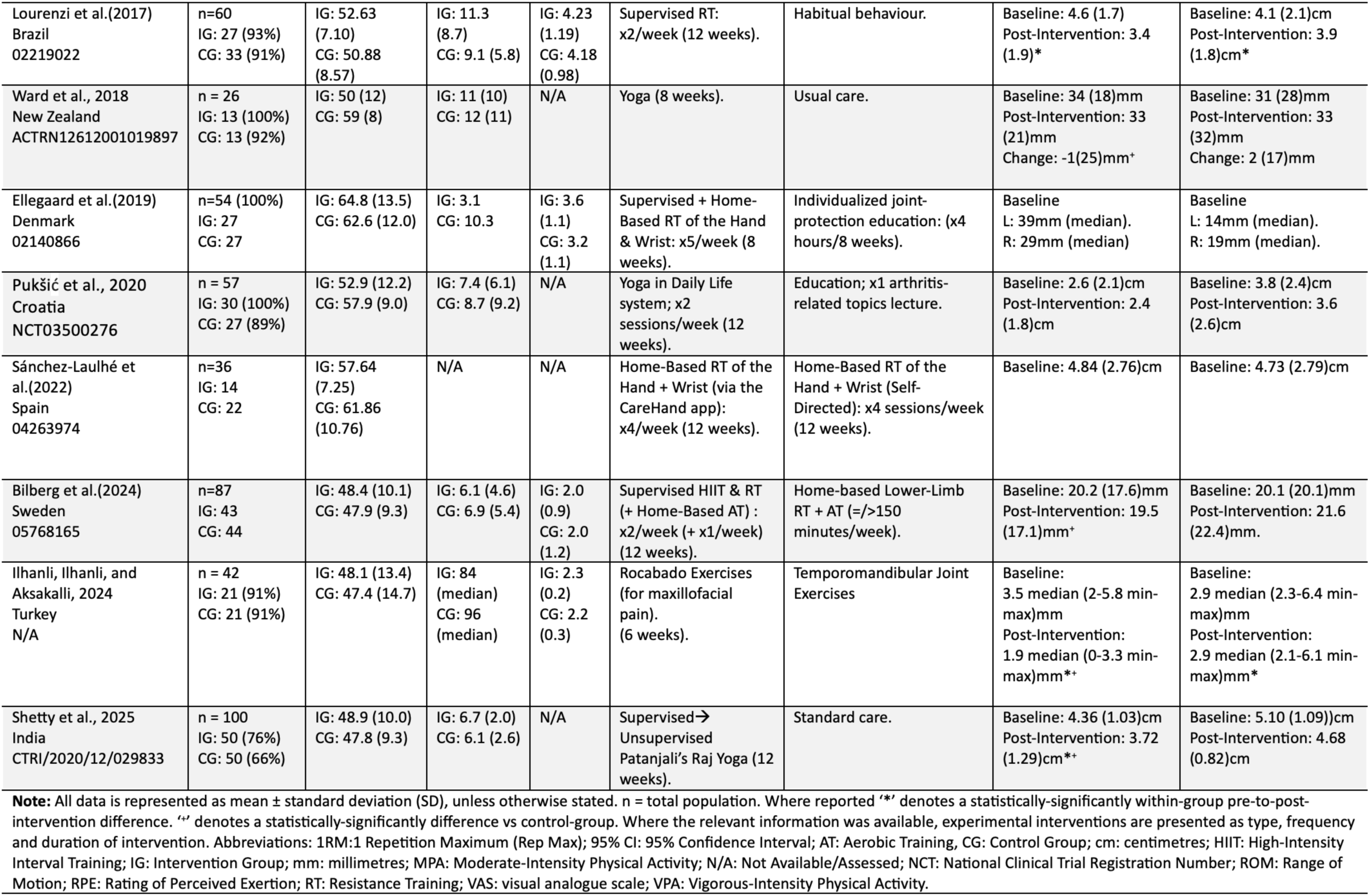
Characteristics of included studies of behavioural interventions. **Alt text:** Table summarizing the key interventional details and participant characteristics of randomized controlled trials of included behavioural interventions.

#### Pharmacological Studies

A total of 11 RCT’s involving 21 monotherapy treatment arms were included within this review (Table 3); hydroxychloroquine (400mg)[74], celecoxib (100mg/200mg/400mg)/naproxen (500mg)[75], leflunomide(<20mg)/sulphasalazine (<2.0g)[76], leflunomide (20mg)/methotrexate (7.5mg)[77], naproxen (500mg)[78], ketoprofen (20mg)[79], tofacitinib (5mg/10mg)[80], baricitinib (2mg/4mg)[81], filgotinib (50mg/100mg/200mg)[82], upadacitinib (15mg/30mg)[83], and sarilumab (200mg)[84]. Studies were conducted within the USA (n=5), Japan, and multi-nationally (n=5), with a duration of either 2, 12 (n=6), 24 (n=3) or 52 weeks. The meta-analysis included 5552 participants across intervention (n=3816) and control groups (n=1736)(69%-94% female); mean age and disease duration ranged from 36-59 years (mean: IG = 54.2, CG = 53.3) and 2.3-14.5 years respectively (mean: IG = 9.0, CG = 8.6).

**Table 3.**
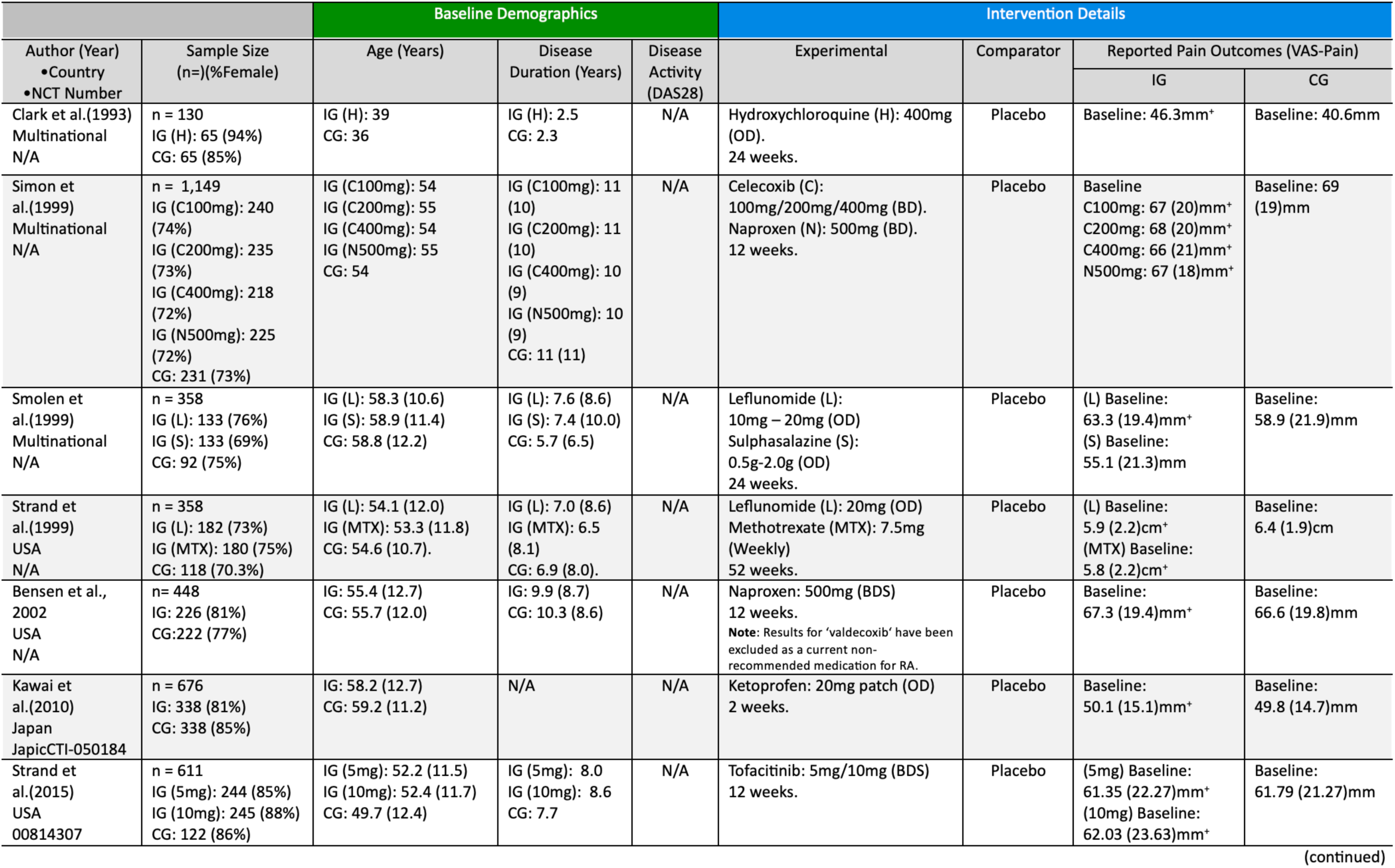
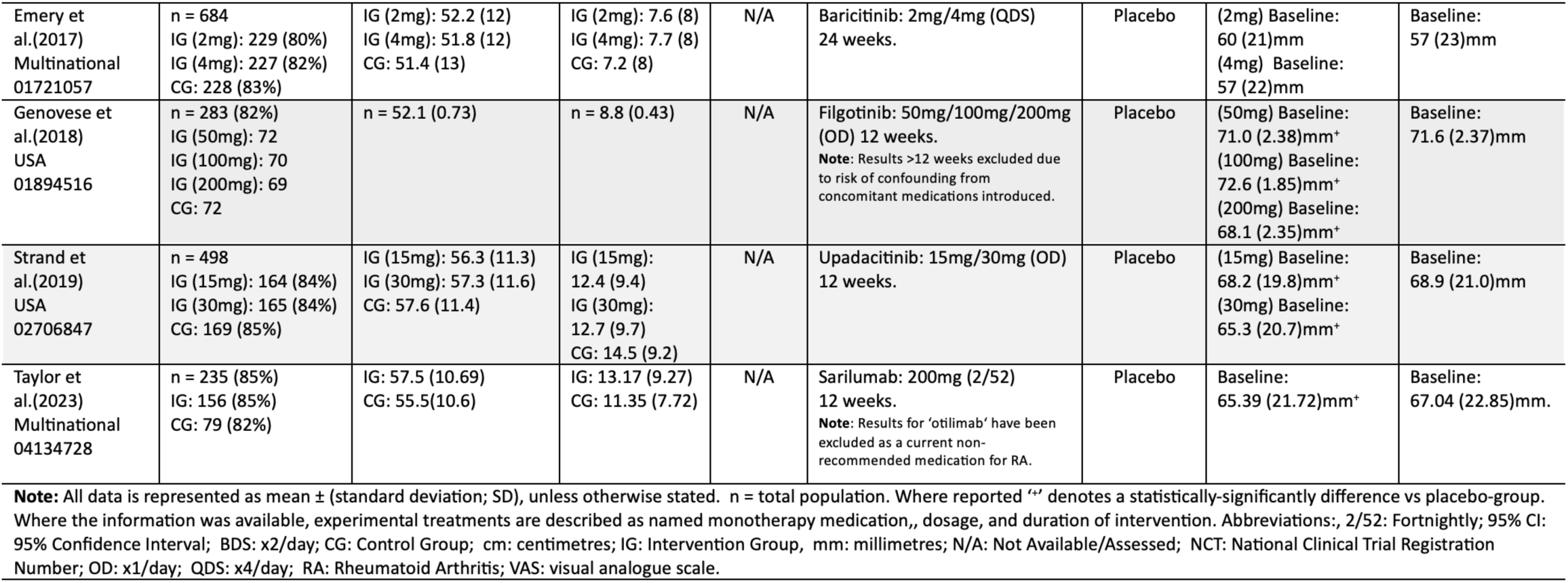
Characteristics of included studies of pharmacological interventions. **Alt text:** Table summarizing the key interventional details and participant characteristics of randomized controlled trials of included pharmacological interventions.

#### Risk of Bias Assessment

In total, of the 25 studies included, four demonstrated low risk of bias; all pharmacological [80, 82, 84], 12 showed reason for some concerns; one pharmacological [74] and eleven behavioural [61–64, 66–70, 71, 72], and nine presented high risk of bias; six pharmacological [75–78, 79, 81] and three behavioural [60, 65 and 73]. A summary of included studies and full results of each study by domain are available in Supplementary Figure S1 (a-c).

### Effects of Interventions

#### Outcomes on Pain Perceptions

As per inclusion criteria, all studies (n=25) measured and reported pain perceptions using the VAS scale. All results for the individual studies and overall syntheses are presented in Figures 2a and 2b. Sensitivity and sub-group analyses were not able to be performed because of inconsistencies between intervention details (i.e., frequency and intensity of PA, limited number of medications within each class, and/or presentation of results according to gender, etc.).

**Figure 2:**
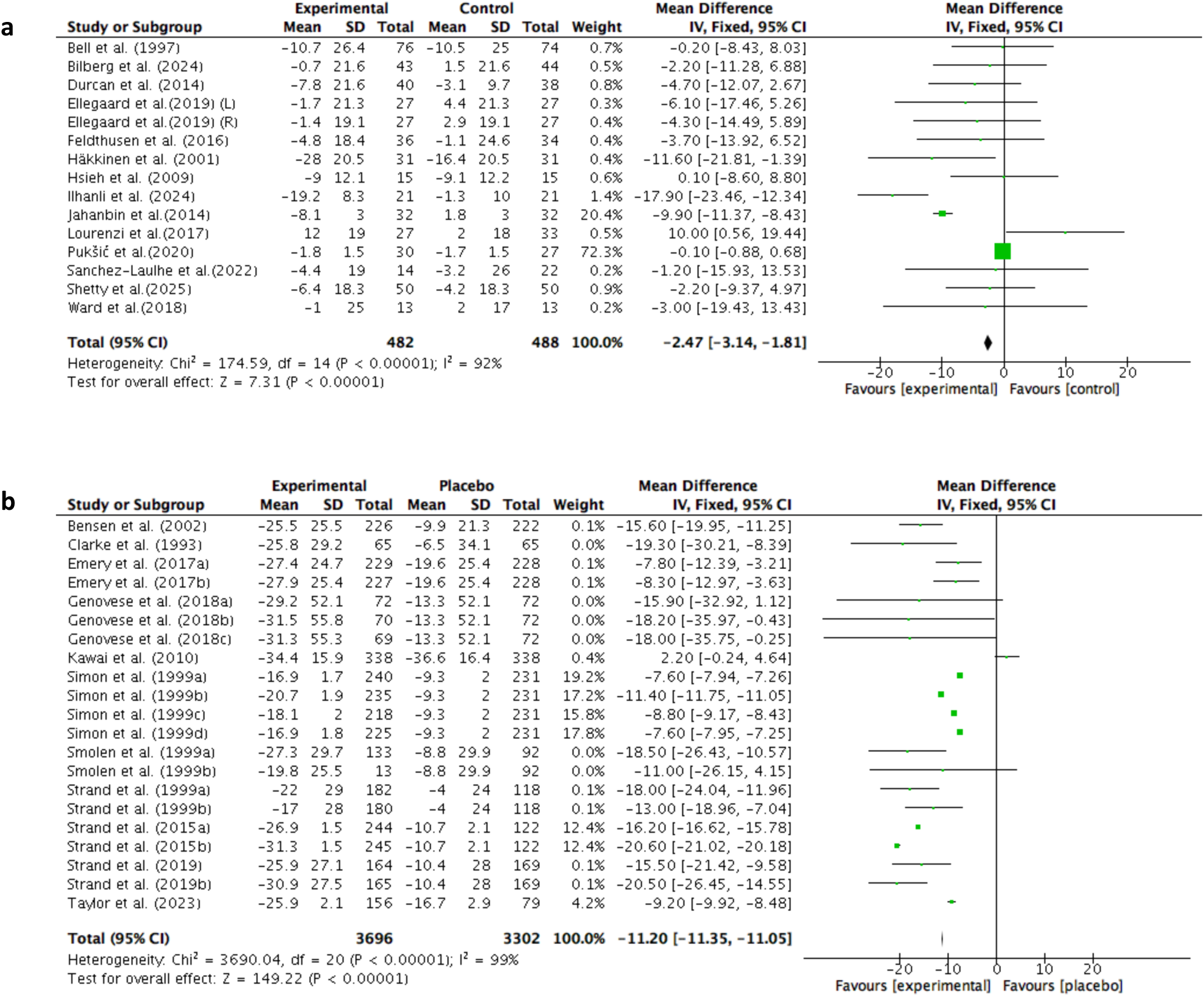
(**a**) Forest plot for changes to VAS-pain scores in response to physical activity interventions. (**b**) Forest plot of changes to VAS-pain scores in response to pharmacological interventions. **Alt text:** Forest plots showing a summary of the mean baseline values and changes/differences in VAS-pain scores of patients with rheumatoid arthritis immediately following the completion of randomized-controlled trials of formally-recommended therapeutic interventions aimed at facilitating pain-management. Separated into (a) behavioural interventions and (b) pharmacological interventions.

### Behavioural Studies

Results of the meta-analysis showed a small, statistically-significant effect of behavioural interventions on pain reduction amongst patients when compared to control groups with a MD of −2.47mm (non-MCID; 95% CI: −3.14 – - 1.81, I^2^ = 92%, z= 7.31, p <0.00001)(Figure 2a). Of the 14 included studies, 10 groups demonstrated a statistically-significant within-group difference for aerobic [61, 62], resistance [65], combined aerobic + resistance training [69], yoga [64], Rocabado exercises [72], and placebo interventions [61 62, 66, 72]. Similarly, seven experimental trials led to a statistically-significant between-group difference in pain-perceptions following programs of aerobic [62], resistance [67], aerobic + resistance training [68, 69], yoga [64, 65], and Rocabado exercises [72].

### Pharmacological Studies

Results of the meta-analysis showed a large, statistically-significant effect of pharmacological interventions on pain reduction in-comparison to placebo-groups with a MD of −11.20mm (MCID; 95% CI: −11.35 – −11.05, I^2^ = 99%, Z = 149.22, p<0.00001) (Figure 2b). Of the 21 intervention arms, all but three medications (sulphasalazine (2.0g)[76] and baricitinib (2mg/4mg) [81]) demonstrated a statistically-significant between-group difference in favour of experimental groups. Unfortunately, it was not possible to compare the significance of within-group differences for experimental or control-groups since this were not reported by any of the included pharmacological studies.

### Certainty of Evidence

Analyses using the GRADE approach demonstrated little influence of varied risk of bias and heterogeneity on the quality of results within this review across included pharmacological studies which were collectively assigned a high certainty of evidence. However, despite an acceptable effect size, due to increased risk of bias, it was determined that included pharmacological studies should be assigned a low certainty of evidence (Table 4).

**Table 4.** Quality of Evidence Assessment. **Alt text:** Table summarizing the factors involved with and completion of individual certainty assessments for the quality of evidence provided by included studies of recommedned behavioural and pharmacological interventions for the management of pain for patients with rheumatoid arthritis

| Summary of findings table according to GRADE analysis |  |  |  |  |  |  | Evaluation components to lower quality |  |  |  |  |  | Evaluative components to higher quality |  |  |
| --- | --- | --- | --- | --- | --- | --- | --- | --- | --- | --- | --- | --- | --- | --- | --- |
| Outcome | Intervention Effects (MD) | No. of studies | No. of participants (IG) | No. of participants (CG) | Grade | Comments | Methodological design start point | Risk of bias | Inconsistency of results | Indirectness | Imprecision | Publication bias | Large effect | Dose response | Confounding |
| VAS-Pain (Be) | MD = - 2.47 (95% CI: -3.14 – -1.81) | 14 | 455 | 461 | Low ⊗⊗ | From the studies including, we are confident that the true interventional effect is close to that of the estimate. | RCTs-only; high-quality. | 79% of studies had 'some concerns' over RoB, and 21% had 'high' RoB; down-grade 1 level. | Heterogeneity can represent inconsistency; I<sup>2</sup> = 92% ('considerable heterogeneity'); downgrade 1 level. | All participants studies met inclusion criteria and PICO strategist line with the research question; no downgrade. | Sample size = 916 and confidence intervals relatively limited; no downgrade. | A comprehensive search strategy was used including scientific databases and best-practice guidance to obtain published and grey literature; no downgrade. | Medium effect size, z=7.31; no upgrade. | Non-present; no upgrade. | No additional factors were found. |
| VAS-Pain (Ph) | MD = -11.20 (95% CI: -11.35 – -11.05) | 11 | 3816 | 1736 | High ⊗⊗⊗⊗ | From the studies including, we are confident that the true interventional effect is close to that of the estimate. | RCTs-only; high-quality. | 55% of studies had 'high' risk of bias; downgrade 1 level. | Heterogeneity can represent inconsistency; I<sup>2</sup> = 99% ('considerable heterogeneity'); downgrade 1 level. | All participants studies met inclusion criteria and PICO strategist line with the research question; no downgrade. | Sample size = 5552 and confidence intervals relatively limited; no downgrade. | A comprehensive search strategy was used including scientific databases and best-practice guidance to obtain published and grey literature; no downgrade. | Large effect size, z= 149.22; upgrade 1 level. | Dose responses present in 5/9 studies; upgrade 1 level. | No additional factors were found. |
| Abbreviations; 95% CI: 95% Confidence Interval; Be: Behavioural Interventions; CG: Control Group; IG: Intervention Group; MD: Mean Difference; Ph: Pharmacological Interventions; PICOS: Participants, Intervention, Comparator, Outcome, Study Design; RCTs: Randomised Controlled Trials; RoB: Risk of Bias. |  |  |  |  |  |  |  |  |  |  |  |  |  |  |  |

Abbreviations; 95% CI: 95% Confidence Interval; Be: Behavioural Interventions; CG: Control Group; IG: Intervention Group; MD: Mean Difference; Ph: Pharmacological Interventions; PICOS: Participants, Intervention, Comparator, Outcome, Study Design; RCTs: Randomised Controlled Trials; RoB: Risk of Bias.

## Discussion

This is a systematic review and meta-analysis of RCTs using the VAS to assess the efficacy of recommended pharmacological or behavioural interventions on the management of pain amongst patients with diagnosed-RA. Overall, data from 25 studies involving 14 PA exercise programs and 21 pharmacological monotherapy arms, published between 1993 and 2025, were synthesised into two meta-analyses to determine the main outcomes of this review. Across 970 participants between groups, yoga, Rocabado exercises, aerobic and/or resistance-training demonstrated a small, statistically-significant effect on pain perceptions versus controls (MD = −2.47mm, p<0.00001). Similarly, amongst participants, individual-use of either NSAIDs, conventional, biological, or targeted-synthetic DMARDs led to a larger, uniform, and statistically-significant difference in pain than that shown by placebos (MD = −11.20mm, p <0.00001). Unsurprisingly, these outcomes are all consistent with the majority of previous reviews and therefore provide invaluable support to one another. A sub-analysis by Ye et al. (2022) [29] found an average reduction of −0.46mm following aerobic and resistance-training programs, supported in a recent literature review by Pham and Lee (2025) [85], albeit of a low-quality of evidence given some uncertainties over interventional details. Through the introduction of aerobic-exercise, Loeppenthin et al. (2022) [86] demonstrated a reduction in pain for some with RA, however within-group confidence intervals of −5.80 – +6.03mm also shows that others experienced a worsening, as found by Eversden et al. (2007) [87]. It is likely that PA had a positive-interaction with pain through acute anti-inflammatory effects by either; increasing micro ribonucleic acids (microRNAs) to supress the activity of pro-inflammatory signalling-molecules (i.e., IL-1, IL-6, etc.) and macrophages [30, 88]; initiating the accumulation of immunoregulatory cytokines (i.e. IL-1 and IL-10) in-response to excessive inflammation [31]; and/or by intensifying the intracranial production of endorphins which can inhibit the descending transmission of pain (a.k.a. exercise-induced hypoalgesia)[88]. In the long-term, PA improves body-composition by promoting the breakdown of adiposity (body fat) that benefits RA-pain since adipokines are known to actively contribute to inflammation [31]. Where PA has failed to influence pain or even been adverse, may have been because of an inappropriate design for the person’s ability (i.e., overexertion) and/or because of inadequacy to fulfil individual needs given heterogeneity in RA-pathology and progression. Treat-to-target strategies (MD: −17.8mm) [90], observational studies which used alternatives to the VAS (n=9) [90], and moderate-to-high quality evidence from RCTs has also previously supported the use of DMARDs for RA-pain having identified greater mean differences in favour of experimental-groups (n=13) [19]. It would be reasonable to assume that the positive effects demonstrated by pharmacological interventions were a direct consequence of their individual immunosuppressive methods-of-action [92; pp.163-251] (as summarized within Supplementary Table S1). Visual analysis of results showed pharmacological interventions to possess a greater therapeutic efficacy on pain; - 15.3mm across studies versus PA interventions. By being biologically-specific, medications can directly-target internal central and peripheral sensory-pathways, attenuating activity of pain mechanisms more immediately. Unfortunately, as per the original rationale of this review, no study had previously chosen to explore this interaction and therefore exists for this outcome to be compared.

To the best of our knowledge, this is the first systematic-review and meta-analysis of RCTs to evaluate and compare the individual therapeutic efficacy of PA and pharmacological interventions for the management of pain within RA. Methods used were comprehensive as they were informed by the Cochrane Handbook for Systematic Reviews of Interventions, and the report written in-accordance with the latest PRISMA guidance (Supplementary Table S2 and S3)[33]. Inclusion criteria according to a PICOS strategy enabled consistency in methods of assessment and participant characteristics, important for generalizability of results. Despite a high-degree of heterogeneity amongst interventions (I^2^ = 99%), GRADE analysis showed meta-analyses for pharmacological interventions to provide a high-quality level of clinical-evidence, possibly explained by the choice to limit studies exclusively to RCTs. Although a large number of studies demonstrated reason for some concern over risk of bias, the majority of these were behavioural interventions (n=11) and were assigned this solely based on the fact of participant’s unavoidable awareness of their allocated intervention which, however unlikely, could have influenced the accuracy of self-reported pain captured by the VAS. In spite of this however, our meta-analyses also has some limitations. Unfortunately, due to unavoidable methodological limitations where trial participants could not have been blinded to behavioural interventions which require autonomous active participation, results of this meta-analyses had to be deemed as a low-level certainty of evidence based on the potential for detection-bias criteria-alone. Inconsistencies in controlling of patient’s previous/background medications may have influenced the magnitude of mean differences amongst pharmacological interventions. Prescription within PA interventions, such as the intensity and total amount of activity, varied greatly across included experimental-groups. Many studies failed to monitor adherence to programs without a consistent method to measure engagement with PA, not taken into account when drawing conclusions. Manual calculations during statistical analysis present a risk of human error in data. PA presents a safe and accessible approach for many patients, where any side effects of this when individualized and/or medications are likely to be more tolerable than coping with uncontrolled-RA. Independently, NSAIDs, conventional, biological, or targeted-synthetic DMARDs, aerobic and/or resistance training appear to play valuable roles in successful pain-management for patients with RA, justifying their inclusion within best-practice guidance. For the majority of patients, adherence to pharmacological interventions can provide greater pain relief, and should therefore be encouraged as a primary intervention within clinical practice for which PA of either modality can provide an evidence-based, but slightly-less effective alternative. Healthcare professionals can feel empowered to use this information to structure and tailor the content of management plans in response to natural variation in RA-presentation. Patients whose pain fails to respond to one intervention can feel reassured that other credible options exist that may better-suited to coping with their individual needs.

As trials continue, future studies should aim to assess the longitudinal, individual therapeutic efficacy and sustainability of benefits from pharmacological (>12 weeks) and PA interventions (>1-year). Effectiveness of PA should be evaluated according to its prescription as the type of activity, intensity (AT: light-vigorous, 50%-95% HRmax, etc; RT: 40%-80% 1RM, 3-8 RPE, etc.), trial-duration (2-52 weeks) and total amount of activity (number x duration of sessions) varied significantly amongst the included studies and therefore was not always clear which aspects were advantageous and would be optimal for clinical practice. Exploring the integrated efficacy of pharmacological and PA interventions is another worthwhile avenue as a combined-approach may present a more comprehensive solution to addressing unfulfilled populational needs. Work is also warranted to help better understand the molecular mechanisms behind anti-inflammatory benefits of aerobic and/or resistance training (i.e., pro-inflammatory cell phenotyping). Future reviews should also aim to address the systematic limitations of this review and expand the search to additional databases.

In conclusion, this review demonstrates high-quality evidence to support the ability of pharmacological and PA interventions to alleviate pain for those with RA. For patients naïve to intervention, individual DMARDs or NSAIDs had a more significant impact on pain-perceptions and therefore appear to be a more-effective initial line of treatment than yoga, or Rocabado exercises, or aerobic and/or resistance training alone.

## Data Availability

All data produced in the present study are presented within the manuscript.
All data obtained from the included studies are contained within the following Microsoft Excel spreadsheet used for formatting during statistical-analysis:
Databases searched were AMED, MEDLINE, CINAHL Plus, SPORTDiscus, EMBASE, Google Scholar and Web of Science. A manual search of reference lists of included studies and NICE evidence reviews for DMARDs and analgesics was also performed to identify potentially-relevant articles.

https://bham-my.sharepoint.com/personal/cxn332_student_bham_ac_uk/_layouts/15/guestaccess.aspx?share=EaIGUYX6nJdAljZWf0KYqrUBZjqzPpkY0tU-4YA_xKO4GA

https://scholar.google.com

https://www.elsevier.com/en-gb/products/embase

https://www.nice.org.uk/guidance/ng100/evidence

## Supplementary data

Supplementary data are available at the following link; Publication Supplementary Materials.pdf

## Abbreviations

1RM: 1 Repetition Maximum (1 Rep Max);
95% CIs: 95% Confidence Intervals;
ACR: American College of Rheumatology;
AT: Aerobic Training;
BDS: Twice-Daily (x2/day);
CG: Control Group;
cm: centimetres;
DMARD: Disease-Modifying Anti-Rheumatic Drug;
EULAR: European Alliance of Associations for Rheumatology;
GRADE: Grading of Recommendations Assessment, Development and Evaluation (GRADE);
HIIT: High-intensity Interval Training;
IG: Intervention Group;
IL-1/6/10/11: Interleukin-1/6/10/11;
JAK/STAT: Janus Kinase/Signal Transducers and Activators of Transcription;
MCID: Minimally Clinically Important Difference;
MD: Mean Difference;
MeSH: Medical Subject Headings;
microRNA: micro ribonucleic acid;
mm: millimetres;
MPA: Moderate-Intensity Physical Activity;
N/A: Not Available/Assessed;
NICE: National Institute of Health and Care Excellence;
NSAID: Non-Steroidal Anti-Inflammatory Drug;
OD: Once-Daily (x1/day);
PA: Physical Activity;
PICOS: Population, Intervention, Comparator, Outcome, Study Design;
PRESS: Peer Review of Electronic Search Strategies;
PRISMA: Preferred Reporting Items for Systematic Review and Meta-Analysis;
QDS: x4/day;
RA: Rheumatoid Arthritis;
RCT: Randomised Controlled Trial;
ROB2: Risk of Bias 2;
ROM: Range of Motion;
RPE: Rating of Perceived Exertion;
RT: Resistance Training;
UK: United Kingdom;
VAS: Visual Analogue Scale;
VPA: Vigorous-Intensity Physical Activity.

## Supplementary Materials

**Supplementary Data S1.**
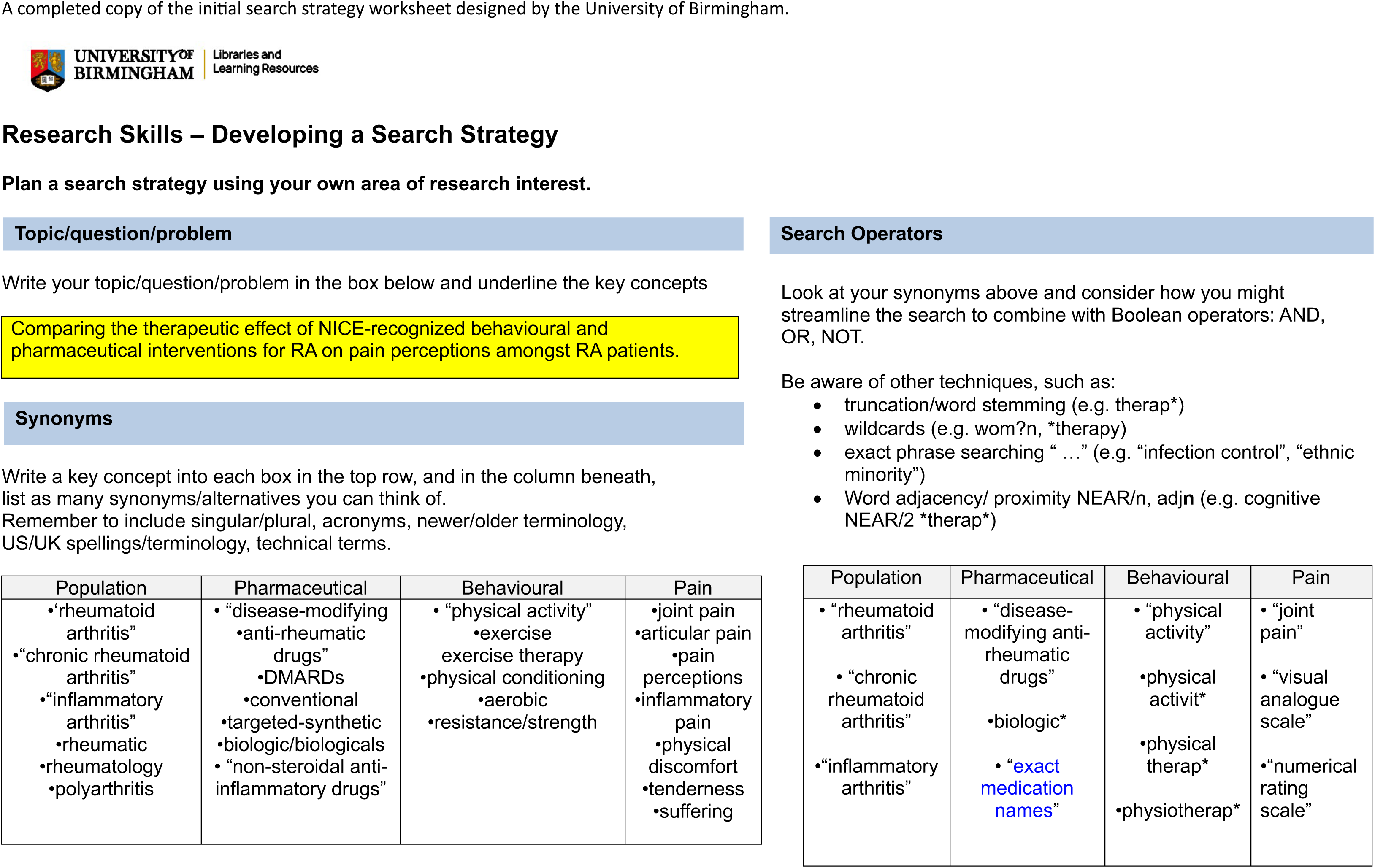
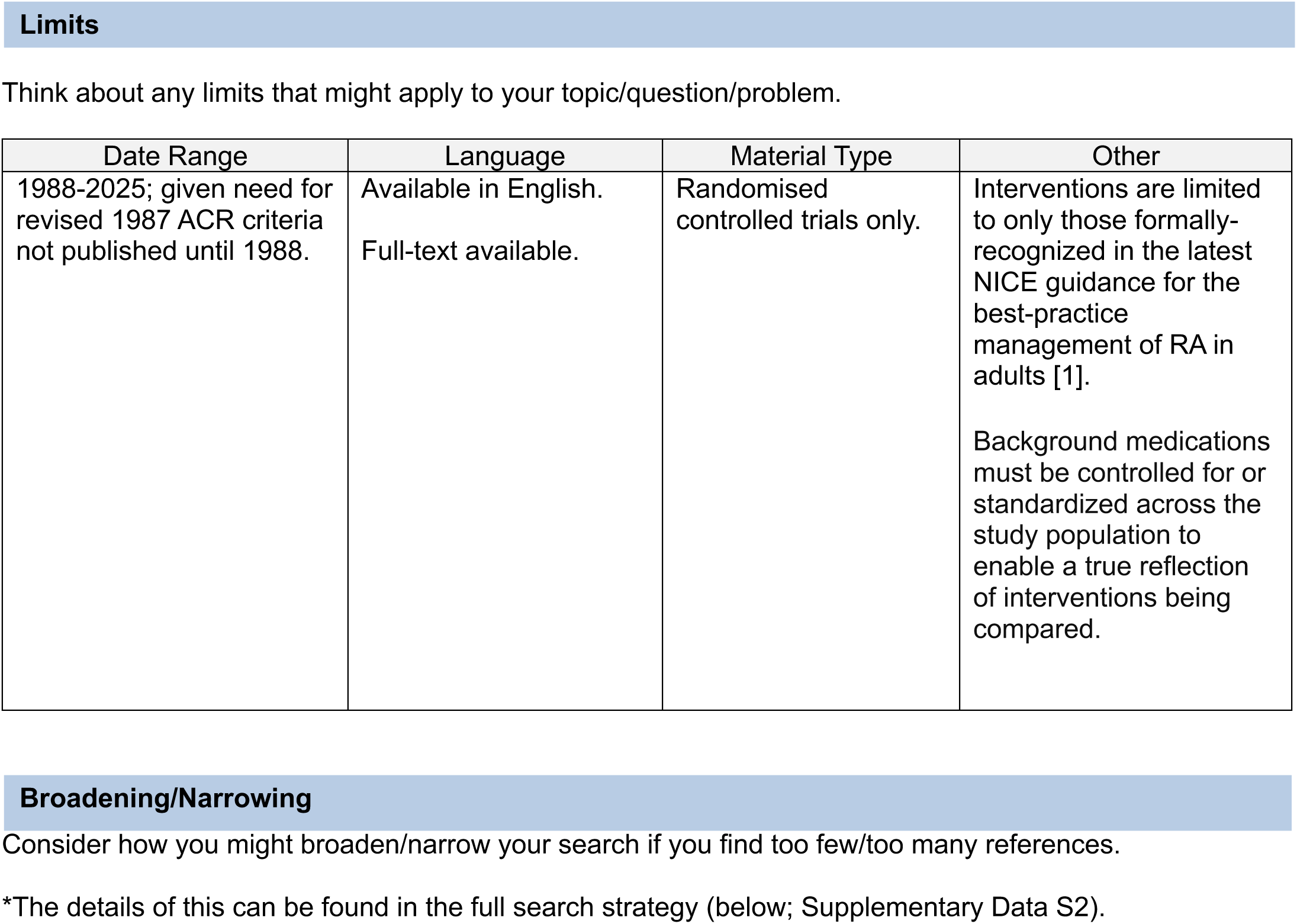

**Supplementary Data S2.**
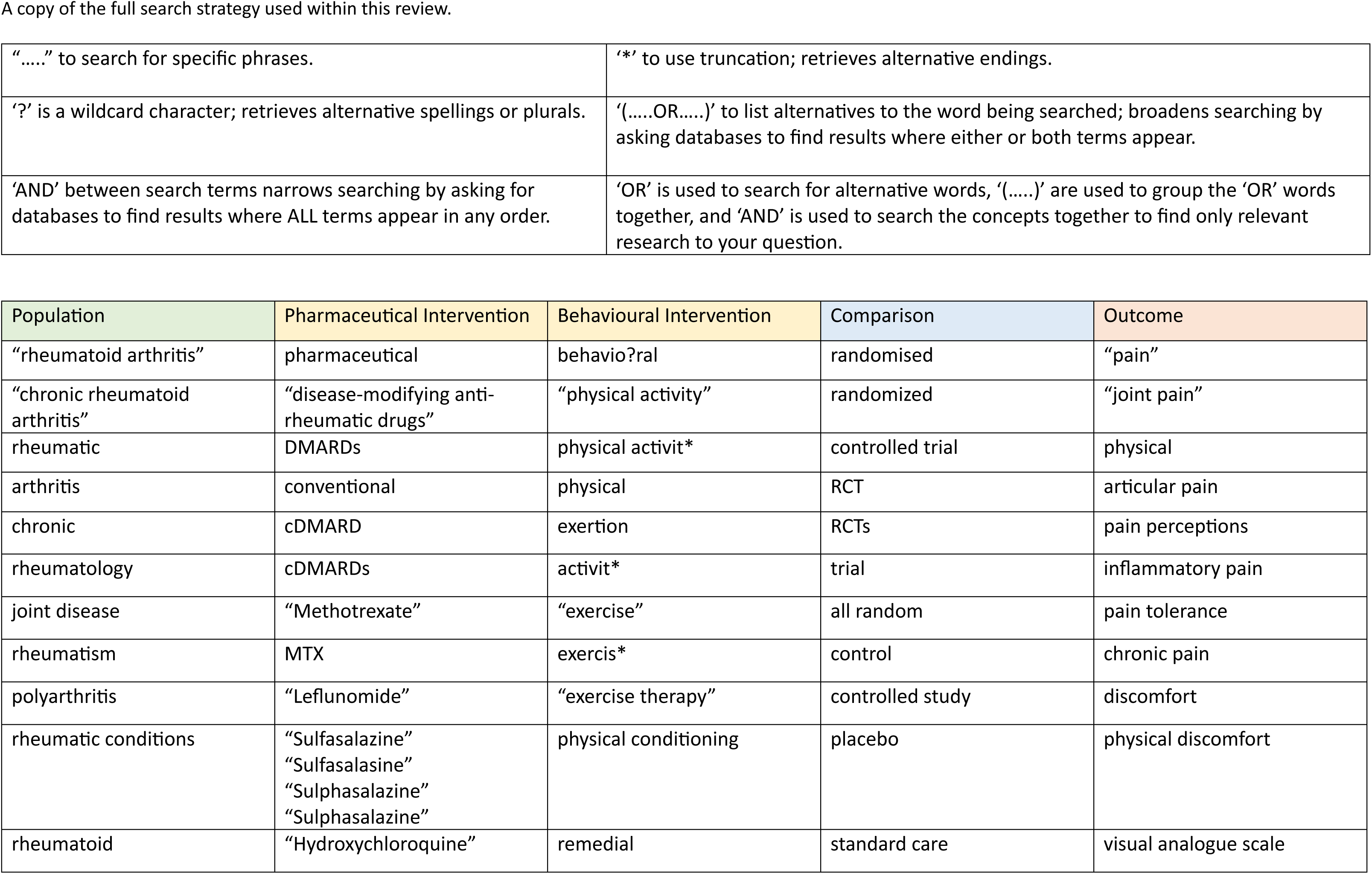
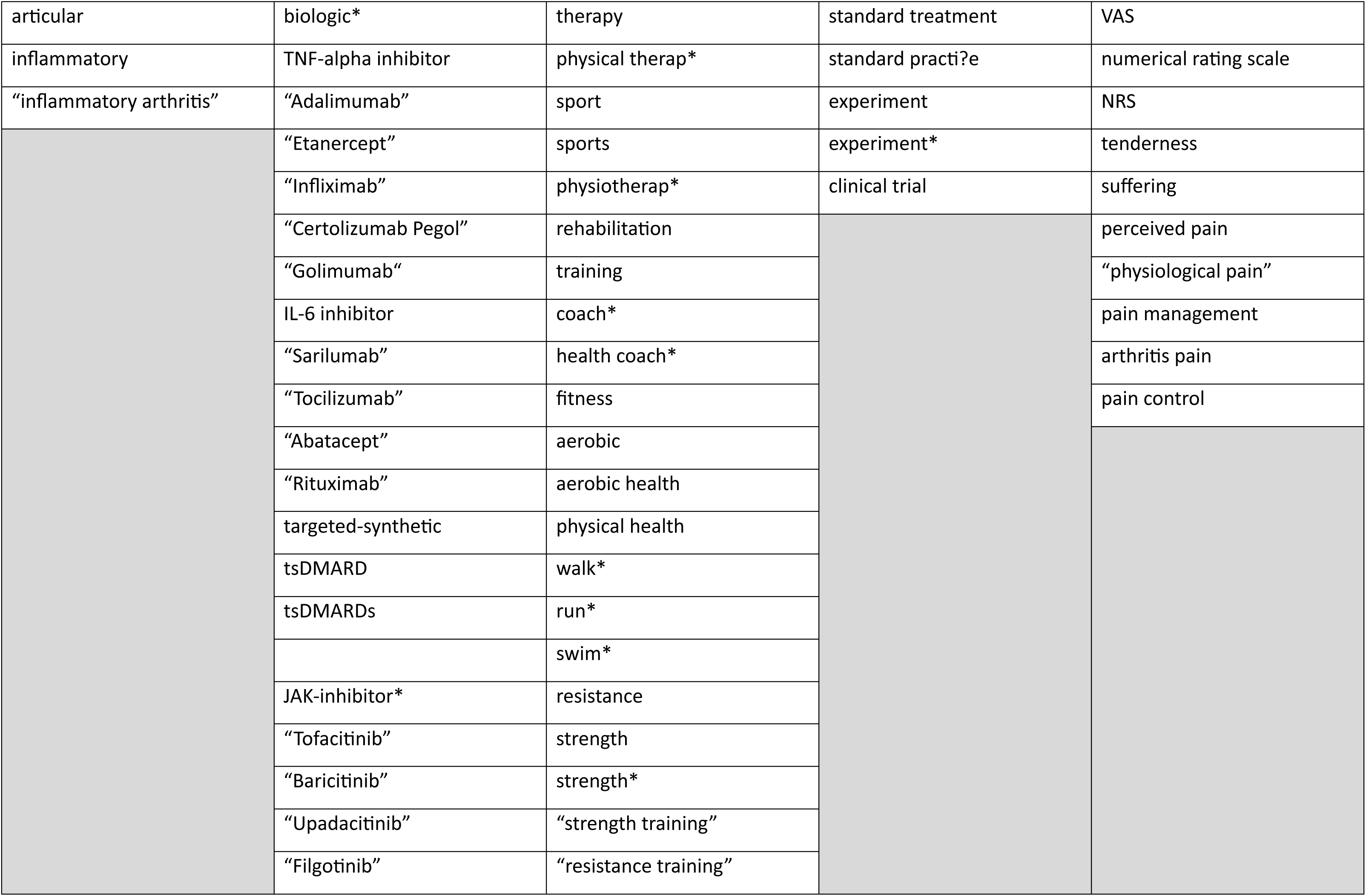
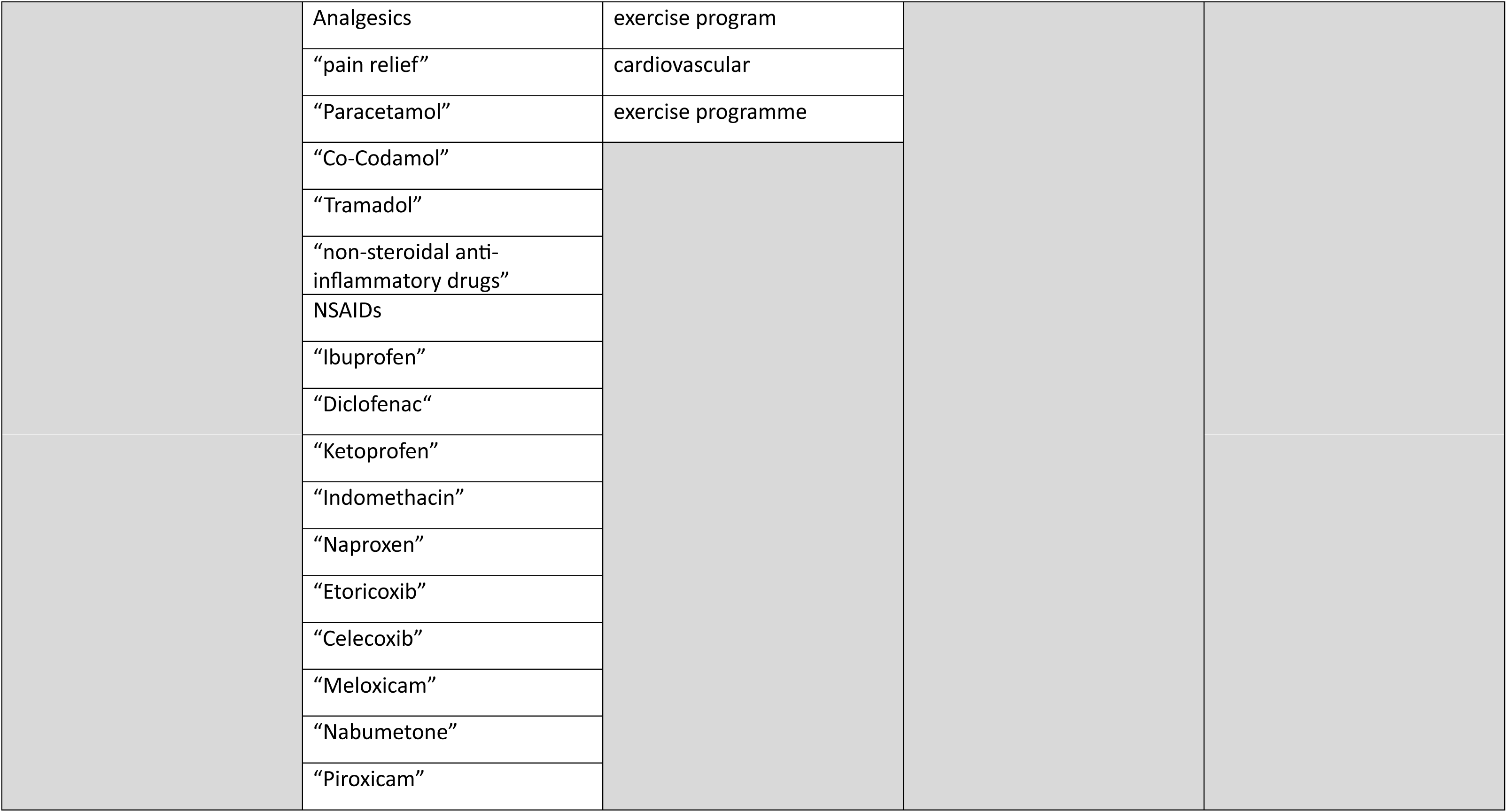
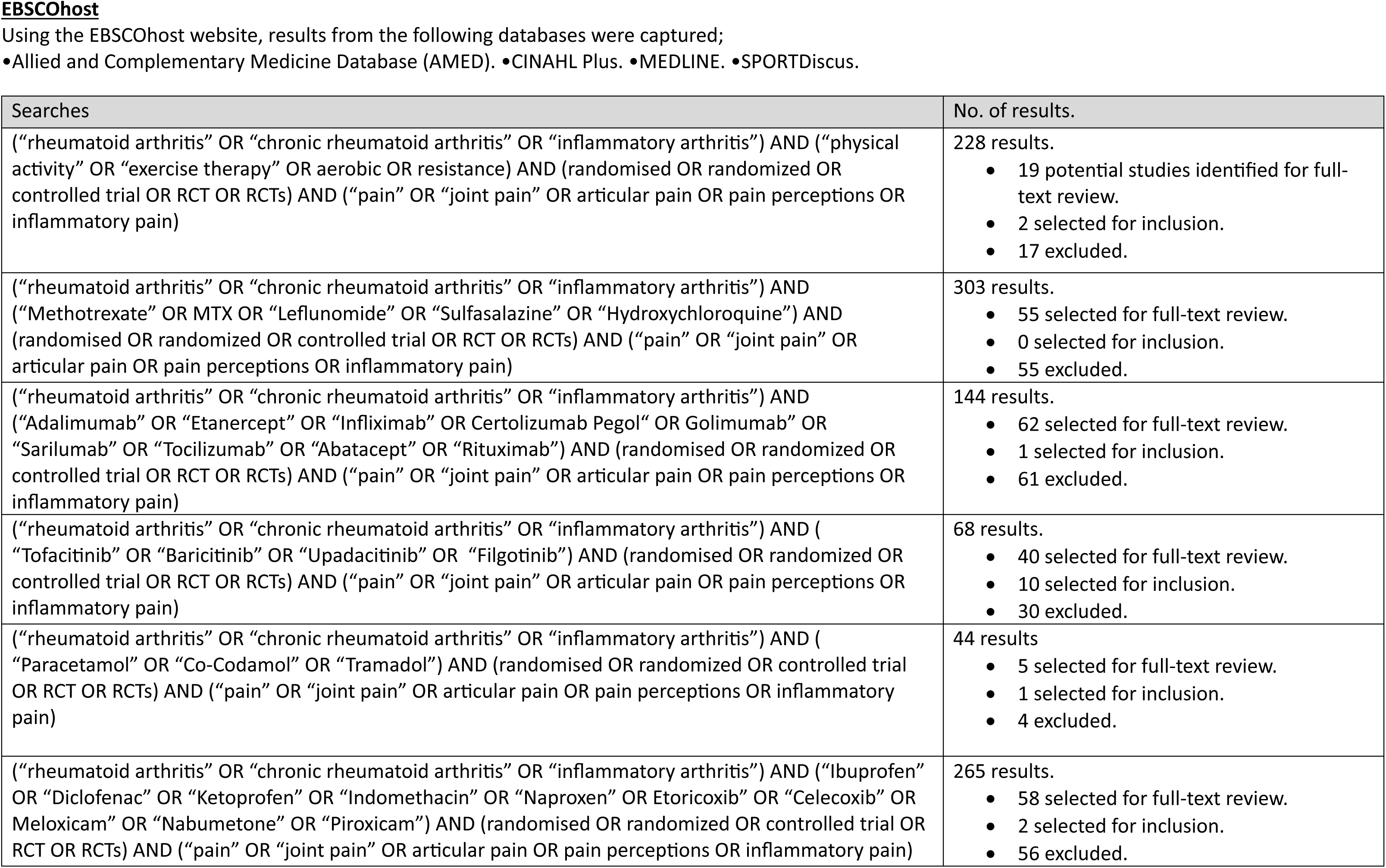
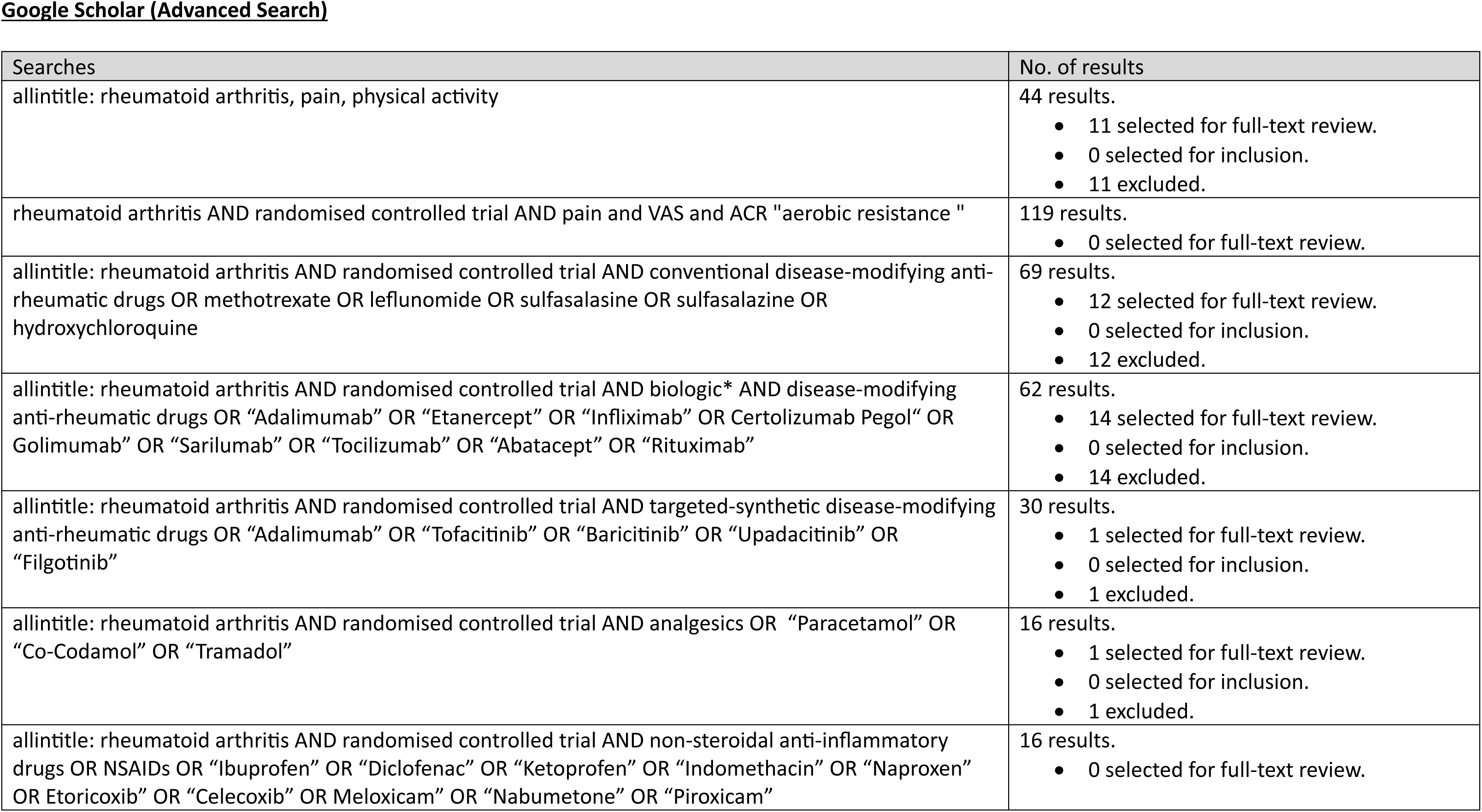
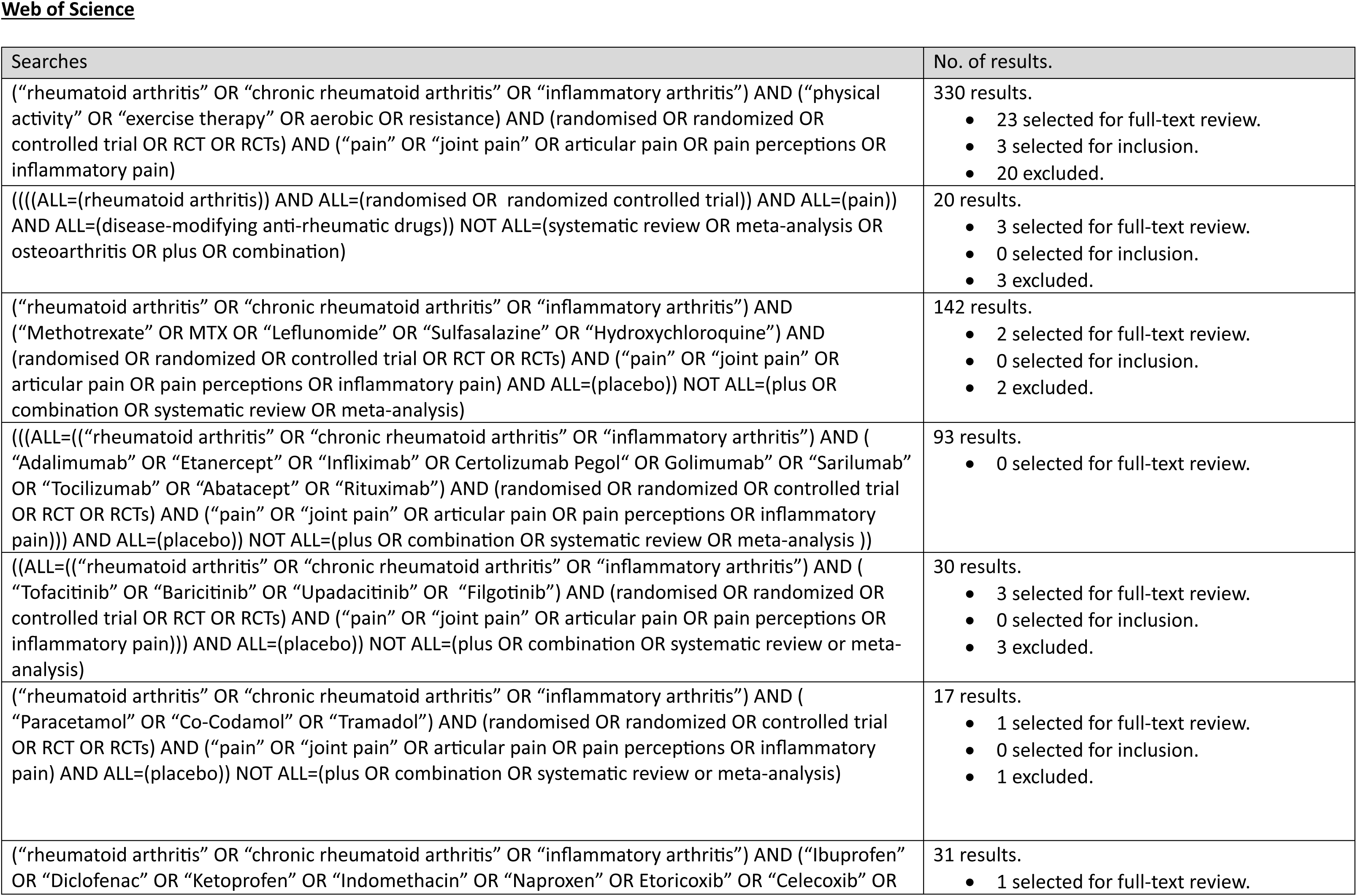
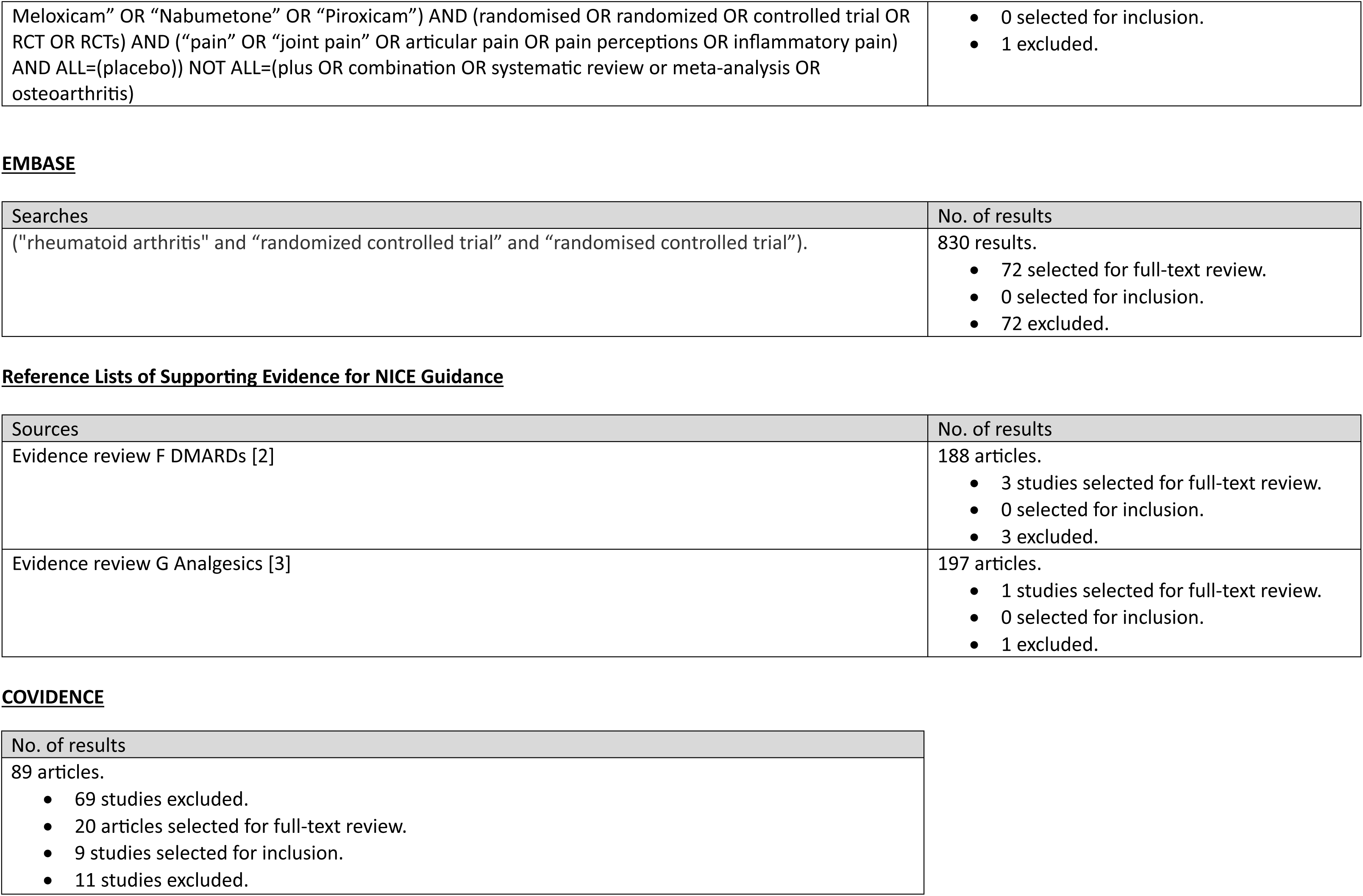

**Supplementary Figure S1.**
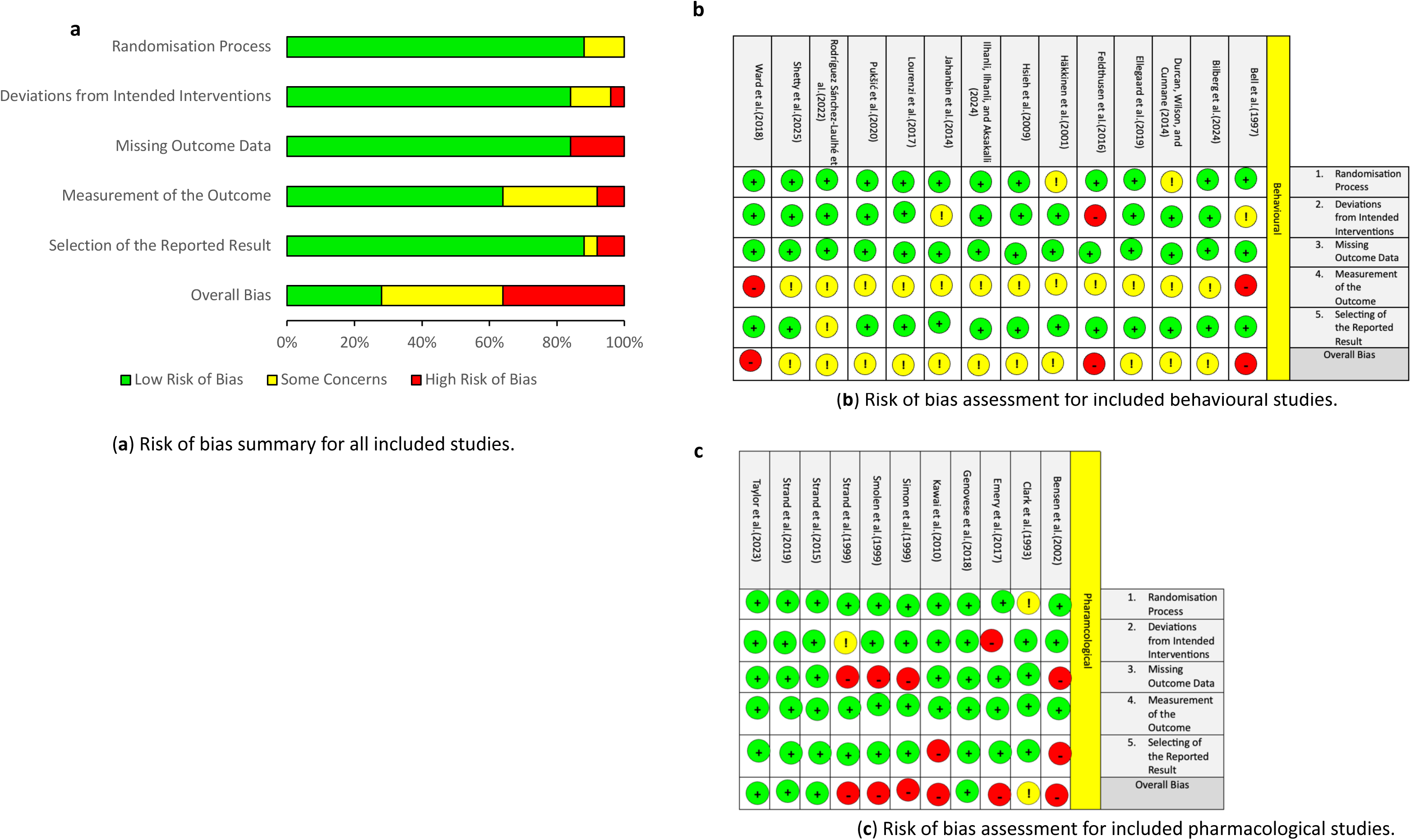
Cochrane-based risk of bias assessment for included studies.

**Supplementary Table S1.** A summary of recommended pharmacological interventions and individual methods of action.

| Medication Class | Method of Action |
| --- | --- |
| cDMARDs | •**Methotrexate**: Increases the concentrations of enzymes responsible for transitioning pro-to-anti-inflammatory macrophages and interferes with DNA synthesis and replication of autoimmune cells [4, 5].<br>•**Sulphasalazine**: Impairs the absorption and transcription of pro-inflammatory macrophages by breaking down folate compounds used for cell division [6].<br>•**Hydroxychloroquine**: Initiates an internal state of alkalosis (increased pH) which opposes antigen-presentation and stops autoimmune-activation of the immune-system [6].<br>•**Leflunomide**: Possesses a mild anti-inflammatory effect and obstructs the synthesis of pyrimidines required for replication of harmful B-/T-lymphocytes [6]. |
| bDMARDs | Provide complementary organic treatment based on biological-compatibility and individual needs within RA pathogenesis [6, 7].<br>•**Adalimumab**: TNF-α Inhibitor. •**Etanercept**: TNF-α Inhibitor.<br>•**Infliximab**: TNF-α Inhibitor. •**Certolizumab Pegol**: TNF-α Inhibitor.<br>•**Golimumab**: TNF-α Inhibitor. •**Sarilumab**: IL-6 Inhibitor.<br>•**Tocilizumab**: IL-6 Inhibitor.<br>•**Abatacept**: Modulator of T-Lymphocyte and Immune Response Activity.<br>•**Rituximab**: Inhibitor of B-Lymphocyte Proliferation [8].<br><u>Key:</u><br>1. A suffix '-cept' refers to a receptor.<br>2. A suffix '-mab' refers a monoclonal antibody [9].<br>3. A suffix '-ximab' refers to a chimeric antibody [9].<br>4. A suffix '-zumab' refers to a humanized antibody; A non-human monoclonal antibody containing a constant region modified to enhance its resemblance of naturally-occurring human variants [10].<br>5. A suffix '-mumab' is a fully-human antibody. |
| tsDMARDs | Genetically-engineered, synthetic small molecules which bind to receptor-sites on pro-inflammatory cytokines in a lock-and-key manner, suppressing the activity of JAK molecules, and thereby preventing intra-cellular signalling of inflammation via the JAK/STAT pathway [11, 12].<br>•**Tofacitinib**: JAK1-3 inhibitor. •**Baricitinib**: JAK1-2 inhibitor. •**Upadacitinib**: JAK1 inhibitor. •**Filgotinib**: JAK1 inhibitor. |
| NSAIDs | Inhibit the activity of cyclooxygenase (COX) 1 and 2 enzymes and prostaglandins, preventing the stimulation and proliferation of pro-inflammatory cells [13]. |
| Analgesics | As per NSAIDs, and also inhibit descending pain transmission [14]. |
| Glucocorticoids | Provide non-specific immunosuppression and symptom-relief as it is a well-known fact that DMARDs take time to act ($\leq 24$ weeks) [6, 15].<br>*These are only to be used as short-term bridging therapy due to well-documented adverse side effects of long-term use. |
| Abbreviations: COX: Cyclooxygenase; DMARDs: Disease-Modifying Anti-Rheumatic Drugs; DNA: Deoxyribonucleic Acid; IL-6: Interleukin-6; JAK: Janus Kinase; NICE: National Institute for Health and Care Excellence; STAT: Signal Transducer and Activator of Transcription; TNF-α: Tumour Necrosis Factor-alpha. | |

**Supplementary Table S2.**
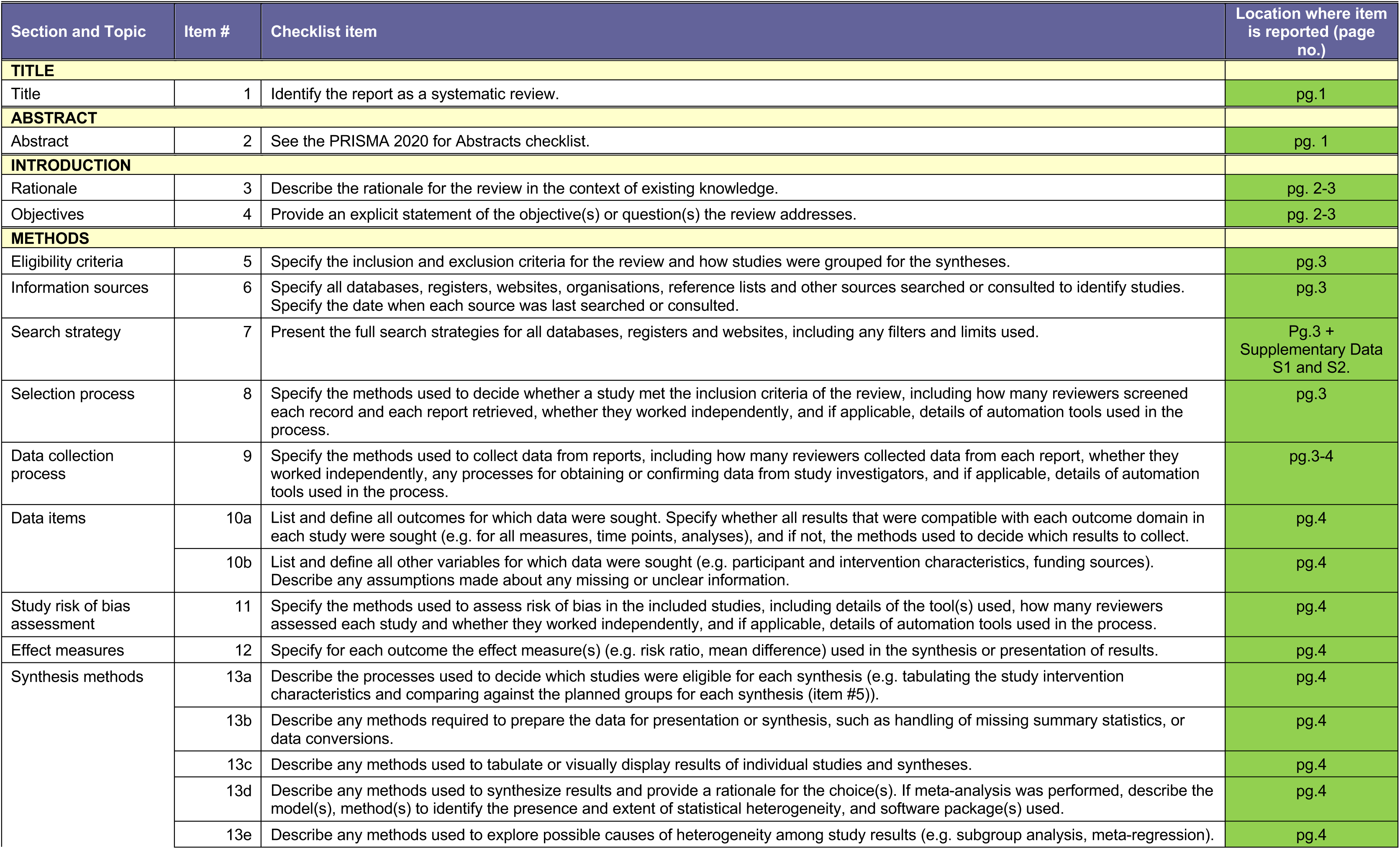
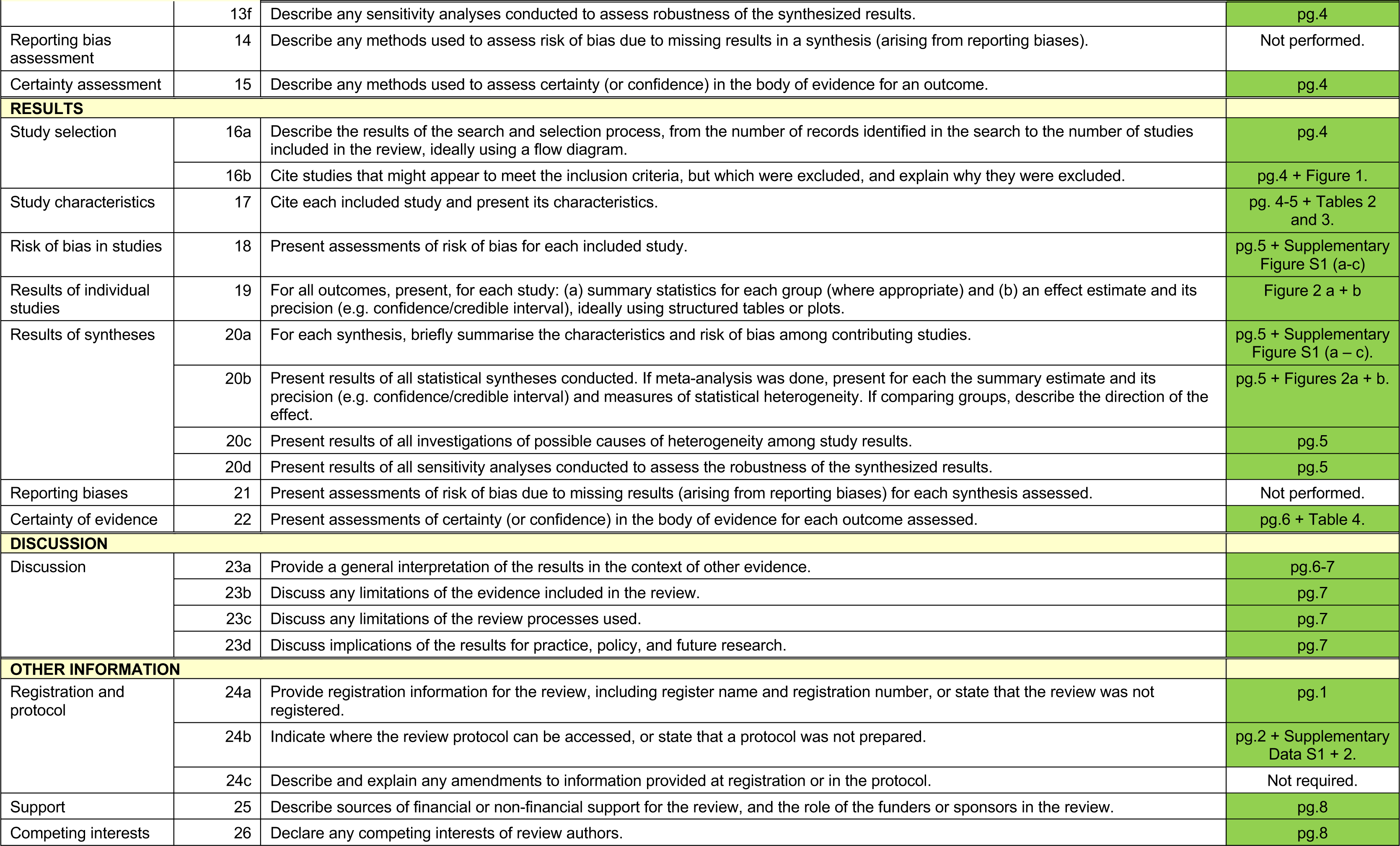
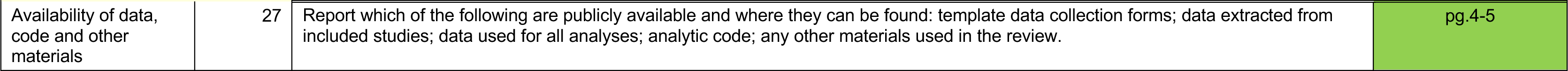
Accordance of report to PRISMA 2020 Full Checklist (adapted from [16])

**Supplementary Table S3.** Accordance of report to PRISMA 2020 Abstract Checklist (adapted from [16])

| Section and Topic | Item # | Checklist item | Reported (Yes/No) |
| --- | --- | --- | --- |
| **TITLE** |  |  |  |
| Title | 1 | Identify the report as a systematic review. | Yes |
| **BACKGROUND** |  |  |  |
| Objectives | 2 | Provide an explicit statement of the main objective(s) or question(s) the review addresses. | Yes |
| **METHODS** |  |  |  |
| Eligibility criteria | 3 | Specify the inclusion and exclusion criteria for the review. | Yes |
| Information sources | 4 | Specify the information sources (e.g. databases, registers) used to identify studies and the date when each was last searched. | Yes |
| Risk of bias | 5 | Specify the methods used to assess risk of bias in the included studies. | Yes |
| Synthesis of results | 6 | Specify the methods used to present and synthesise results. | Yes |
| **RESULTS** |  |  |  |
| Included studies | 7 | Give the total number of included studies and participants and summarise relevant characteristics of studies. | Yes |
| Synthesis of results | 8 | Present results for main outcomes, preferably indicating the number of included studies and participants for each. If meta-analysis was done, report the summary estimate and confidence/credible interval. If comparing groups, indicate the direction of the effect (i.e. which group is favoured). | Yes |
| **DISCUSSION** |  |  |  |
| Limitations of evidence | 9 | Provide a brief summary of the limitations of the evidence included in the review (e.g. study risk of bias, inconsistency and imprecision). | Yes |
| Interpretation | 10 | Provide a general interpretation of the results and important implications. | Yes |
| **OTHER** |  |  |  |
| Funding | 11 | Specify the primary source of funding for the review. | Not required. |
| Registration | 12 | Provide the register name and registration number. | PROSPERO<br>CRD420251069339 |

